# From amplicon to antigen: a quantified transmission map that nominates multi-antigen antibody–drug-conjugate co-target sets across cancer types

**DOI:** 10.64898/2026.07.13.26357987

**Authors:** Jie Min Lam, Simon Walker-Samuel, Adam Pennycuick

## Abstract

Somatic copy-number amplification is pervasive in cancer, and the genes it carries are candidate drug targets—but only those whose amplification is transmitted to accessible surface protein can be reached by an antibody–drug conjugate (ADC). We build an integrated map of copy-number-to-protein transmission across six tumour types and ask, for every amplified gene, whether its dosage reaches the surface. Copy number transmits to mRNA (median per-gene r = 0.21) but is attenuated at the protein level in 85% of genes, and the mRNA ranking is largely preserved to protein (*ρ* = 0.70); the ranking is set principally at the chromatin/transcription step—among directly measured regulatory inputs, promoter DNA methylation and tumour chromatin accessibility each explain about an order of magnitude more of the transmission variance than gene structure, and do so complementarily. Critically, transmissibility is a stable, gene-intrinsic property: it is predictable from gene properties alone, with no proteomic input, at a leave-gene-out rank correlation of 0.52 (R^2^ = 0.29); it is not positional (holding out whole chromosome arms changes accuracy by 0.001); and it transfers across lineages (Kendall W = 0.97 across leave-one-lineage-out refits). This licenses a predictor that nominates surface targets in cancer types that lack a tissue-referenced proteome, combining direct protein measurement where it is available with prediction where it is not. Requiring co-elevation on a recurrent amplicon with measured transmissibility and an accessible extracellular ectodomain nominates 22 surface antigens on 18 distinct recurrent amplicons across four cancer types (renal, endometrial and both lung subtypes)—for example ITGB8+TSPAN13+TTYH3 on lung 7p, NCSTN+HSD17B7+MPZL1 on 1q (recurrent in several types), the transferrin receptor TFRC on squamous 3q, and FZD1 on clear-cell renal 7q; 21 of the 22 are non-driver passengers and 10 are confirmed on the experimental Cell Surface Protein Atlas. In single malignant cells, against a null that controls for per-cell sequencing depth, the co-detected constructs sit at a modest 1.05–1.45× above independence (p < 0.001, donor-block bootstrap intervals clear of 1.0), and at binding-relevant thresholds the normal-tissue co-expression collapses—so an avidity AND-gate that binds stably only where the antigens co-occur would spare normal cells that carry only one. Observed transmissibility itself transfers strongly between the two lung subtypes (*ρ* = 0.88) and remains positive across distant lineages, consistent with the shared cell-of-origin regulation the map implies. Single-cell co-detection is demonstrated wherever a malignant single-cell atlas exists (both lung subtypes and glioblastoma — the latter entirely from prediction, using no GBM surface-abundance measurement); the remaining cohorts are nominated on the same genetic and topological evidence. The result is a pan-cancer, confidence-tiered catalogue of multi-antigen ADC co-target sets with a concrete plan to test them.

## Introduction

Most solid tumours carry large somatic copy-number amplifications, and these segmental gains are among the earliest and most consistent events in carcinogenesis(11–13). Some recur so reliably in a given cancer type—3q in lung squamous carcinoma is the textbook case(4, 5)—that the gain is thought to be selected for, and therefore present in the great majority of tumours of that type. An amplicon spans many genes at once. A few are the oncogenic drivers the tumour is selected to gain; most are innocent bystanders, co-amplified passengers with no role in the cancer’s biology. Those passengers are usually ignored, but they share a valuable property with the driver: because they sit on the same selected-for DNA segment, they are amplified in nearly every tumour that carries the amplicon.

That property matters for a specific class of drug. An antibody–drug conjugate (ADC) is a targeting antibody carrying a cytotoxic payload; it kills any cell that displays enough of its target antigen on the surface, whether or not that antigen does anything for the tumour(1, 2). An ADC therefore does not need to hit a driver—it needs a surface protein that is abundant on tumour cells and scarce on normal ones. A co-amplified passenger displayed on the surface is exactly such a target. And because an amplicon elevates several surface passengers together, a single tumour can present several co-amplified antigens at once, which opens a design a single-target ADC cannot use: a multispecific binder engaging two or three of them simultaneously. The clinical feasibility of multispecific ADCs is now established—a bispecific EGFR/HER3 ADC has reported phase-3 activity(3)— but they have been built against pairs of driver receptors, not against sets of co-amplified passengers. Targeting a passenger *set* defined by one amplification is, to our knowledge, a new idea, and it offers two advantages at once: engaging several antigens sharpens selectivity for cells that carry the whole amplicon, and requiring several targets makes resistance by antigen loss harder to evolve, because escaping the drug would mean losing a segment the tumour is selected to keep.

The obstacle is that an amplification is not a surface protein. A gene gained at the DNA level need not rise at the mRNA level, and a gene raised at the mRNA level need not reach the protein or the cell surface: copy-number dosage is heavily attenuated on the way to protein, unevenly from gene to gene(6–10). Which co-amplified passengers actually reach the surface—and can therefore be reached back by an antibody—is not obvious from the genome. This paper builds a quantified map of that amplicon-to-surface path, uses it to nominate multi-antigen ADC co-target sets across six cancer types, and lays out a staged experiment to test them.

## Results

### The transmission cascade and where it is gated

The analysis draws on three nested tiers of data, and it helps to state them up front. All six CPTAC cohorts have matched copy number, mRNA and raw protein and so enter the transmission cascade; four of the six additionally have a proteome referenced to matched normal tissue (CCRCC, LSCC, LUAD, UCEC) and so support measured surface-target nomination; and three cancer types have a malignant single-cell atlas (LUAD, LSCC and GBM) in which same-cell co-detection can be tested. Cohorts therefore drop out at successive steps not arbitrarily but because a specific data type is absent, and each transition is noted where it occurs.

We first quantify the cascade from amplicon to surface. For each gene we define two quantities across six CPTAC cohorts with matched copy number, mRNA and protein (CCRCC, GBM, LSCC, LUAD, PDA and UCEC; 110, 98, 108, 105, 136 and 99 tumours with all three layers jointly measured): copy-number-to-mRNA transmission (the Pearson correlation, across tumours, between a gene’s copy number and its mRNA) and copy-number-to-protein responsiveness (the same correlation to protein). Their difference is the post-transcriptional attenuation. Three related terms recur below and are worth separating at the outset: *transmission* and *responsiveness* are these two per-gene correlations, *attenuation* is their difference, and *transmissibility* (defined in the next section) is a distinct quantity—not a correlation but the fraction of amplified tumours in which a gene’s protein is actually elevated—which is what the predictor learns. Correlations are computed per gene and per cancer type on jointly non-missing cases (minimum 20 cases per type, at least two types) and averaged across types by Fisher-z weighting on n-3 (Supplementary Methods). Across the 6,979 genes meeting this support, copy number transmits to mRNA with a median per-gene correlation of 0.21 but only 0.11 to protein, a median attenuation of 0.08 that leaves 85% of genes attenuated at the protein level (Figure 1a,b). Crucially, the between-gene ranking established at the mRNA step is largely preserved to protein (Spearman *ρ* = 0.70, n = 6,979 genes): the genes that transmit strongly to mRNA are, by and large, the same genes that reach high protein. This is what licenses an mRNA- and gene-property-based predictor to stand in for protein measurement.

**Fig. 1.**
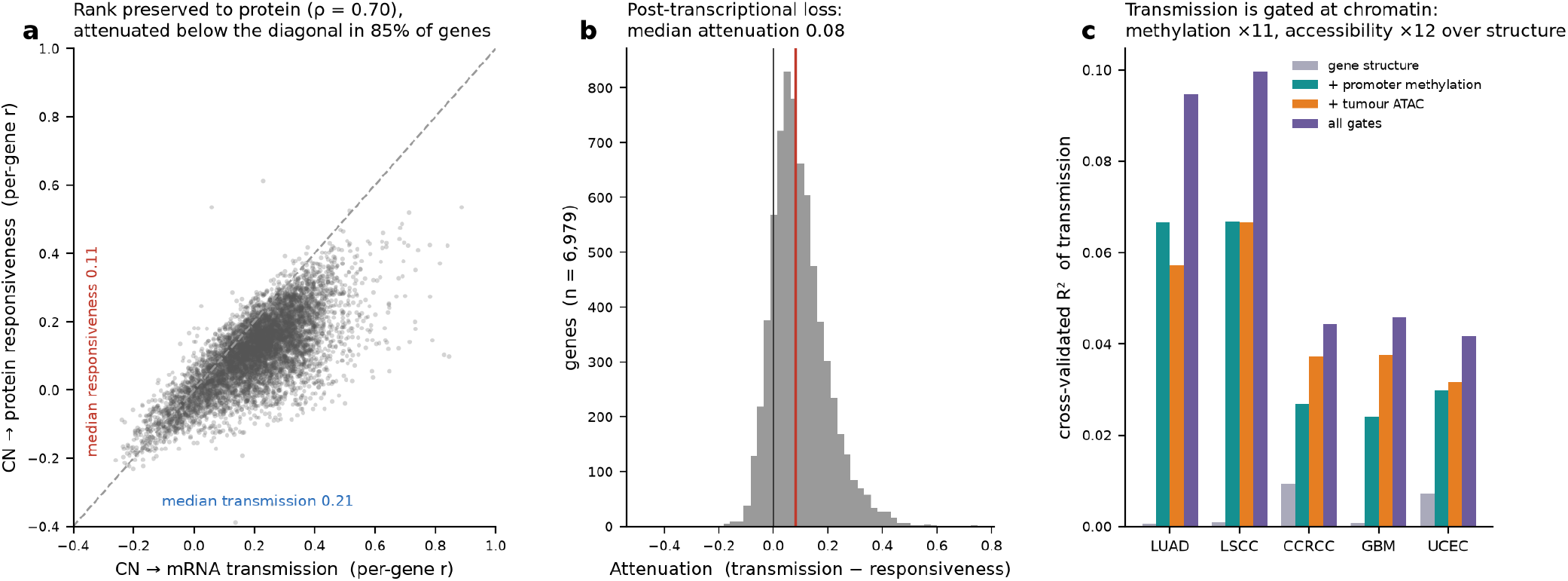
The transmission cascade and its gates, computed on the source-assembled copy-number, mRNA and protein layers (CPTAC; per-gene Pearson correlations across tumours, ≥ 20 cases per cancer type and ≥ 2 types, Fisher-z averaged on n-3). (a) Per-gene copy-number-to-mRNA transmission versus copy-number-to-protein responsiveness (n = 6,979 genes); most genes fall below the diagonal (attenuated post-transcriptionally), and the mRNA-level ranking is preserved to protein (Spearman *ρ* = 0.70, n = 6,979). (b) Distribution of per-gene attenuation (transmission minus responsiveness), median 0.08, n = 6,979 genes. (c) Cross-validated R^2^ of per-gene transmission predicted from gene structure alone, then adding promoter DNA methylation, tumour promoter chromatin accessibility, and both, one group of bars per cancer type (five profiled types: LUAD, LSCC, CCRCC, GBM, UCEC; *≈*4,044 genes per type with matched regulatory data; nested cross-validation). Promoter methylation and accessibility each raise the explained variance about an order of magnitude over structure and are complementary—together reaching mean R^2^ 0.065 versus 0.004 for structure alone.

We then ask where the transmission ranking is set. Gene-structural features alone (local gene density) explain almost none of the between-gene variance in transmission (crossvalidated R^2^ = 0.004, mean over five cancer types with matched regulatory data; ∼4,044 genes per type). Two regulatory layers each add an order of magnitude: adding promoter DNA methylation raises the explained variance to R^2^ = 0.043 ( ≈11-fold over structure), and adding tumour promoter chromatin accessibility raises it to R^2^ = 0.046 (≈12-fold). The two are complementary, not redundant: combining methylation and accessibility reaches R^2^ = 0.065, above either alone (Figure 1c), and the pattern holds across all five profiled cancer types (LUAD, LSCC, CCRCC, GBM, UCEC). Among the regulatory inputs we can measure directly, then, chromatin state dominates gene structure by roughly an order of magnitude. These tumour-measured chromatin features explain only a modest share of the between-gene variance in absolute terms; a broader set of static gene properties, assembled in the next section, predicts a gene’s transmissibility far more completely (R^2^ = 0.32, Figure 2). The two analyses are consistent rather than competing: Figure 1c isolates how much two specific tumour-measured chromatin covariates add over gene structure for one outcome (the copynumber-to-protein correlation), whereas the predictor in Figure 2 pools many gene-intrinsic properties—several of which are themselves upstream determinants of chromatin state—to predict a different, more stable outcome (the transmissibility fraction). The consistent reading is that the transmission ranking is set at the chromatin/transcription step, and that a gene’s own biology, being what shapes that step, is what makes the outcome predictable.

**Fig. 2.**
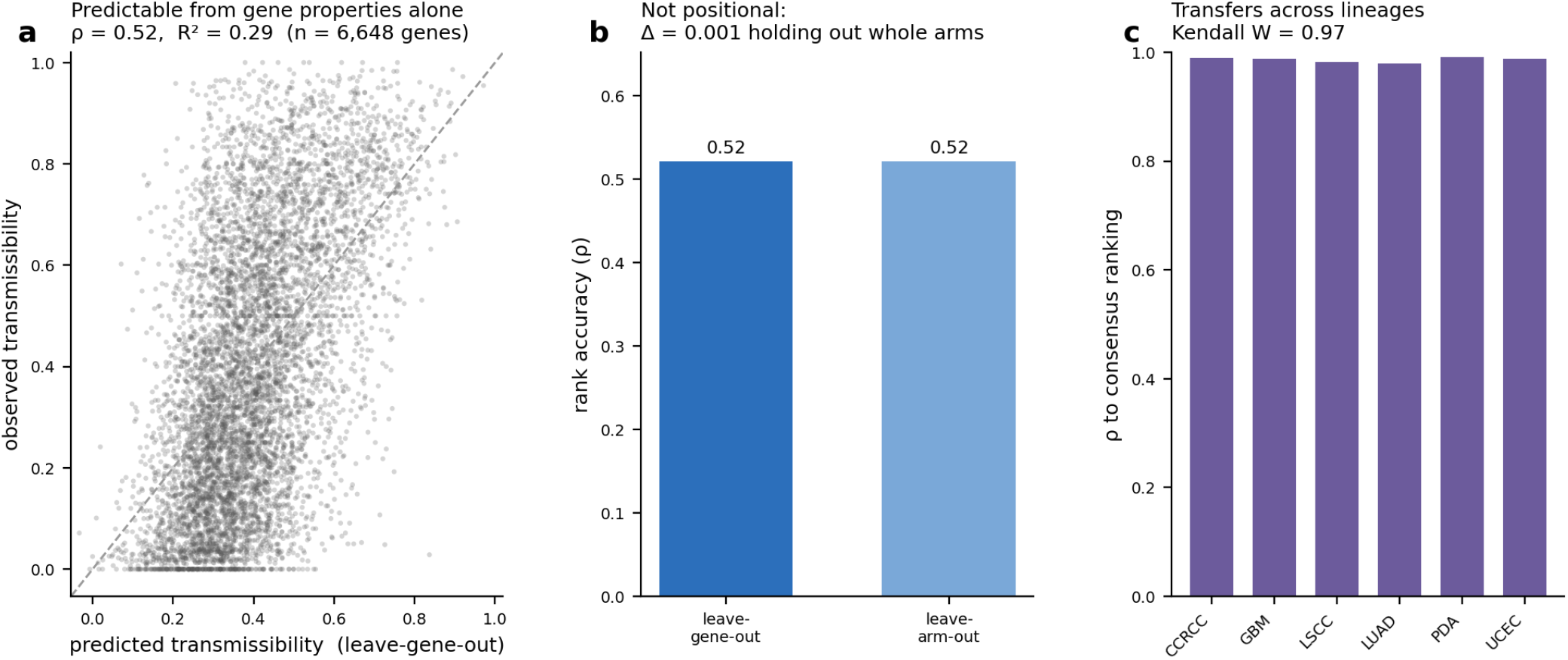
Transmissibility is predictable from gene properties alone (gradient-boosted regressor, 39 gene-property features, no protein-derived input; n = 6,648 genes). (a) Fivefold leave-gene-out out-of-fold predicted versus observed transmissibility (Spearman *ρ* = 0.52, R^2^ = 0.29). (b) Positional control: leave-gene-out versus leave-chromosomearm-out out-of-fold accuracy (Δ*ρ* = 0.001), showing the signal is not carried by same-arm neighbours. (c) Cross-lineage transfer: agreement of predicted rankings across six leave-one-lineage-out refits (Kendall’s W = 0.97, six cancer types).

### Transmissibility is predictable without protein data

We now summarise a gene’s tendency to reach protein by a single scalar, its transmissibility: the fraction of amplified tumours (copy number above the ploidy-adjusted amplification threshold, deduplicated to one value per case) in which the gene’s protein is elevated relative to matched normal tissue. Here “elevated” means a tissue-referenced protein rank above 0.80—that is, the protein sits in the top fifth of its abundance distribution when each tumour’s protein level is ranked against the same protein in the matched normal tissue of origin, so the measure reflects tumour-specific gain rather than a protein that is simply abundant everywhere. Transmissibility runs from 0 (never elevated when amplified) to 1 (always elevated); it is defined per gene, pooled across the cohorts in which the gene is recurrently amplified, and is the quantity the predictor learns.

If transmissibility is an intrinsic property of a gene, it should be predictable from the gene’s own biology, measured independently of any proteomics. Using only gene properties—dosage sensitivity, protein biophysics, mRNA features, evolutionary constraint, complex membership, expression breadth and network centrality, each defined with its data source in Supplementary Table S1(16–18)—and a leave-gene-out design in which no gene informs its own prediction, we recover transmissibility at a rank correlation of 0.52 (R^2^ = 0.29) across 6,648 genes (a marginally smaller set than the 6,979 of the cascade, because a gene enters here only if it has both a defined transmissibility and a complete gene-property feature vector). No protein-derived feature enters the predictor. Two controls establish that this is intrinsic rather than positional or lineage-bound. Holding out whole chromosome arms in turn, so no gene is predicted from a same-arm neighbour, changes the accuracy by only 0.001—the signal is not a positional artefact. And refitting the predictor with each lineage held out in turn—so a cohort’s own tumours never inform the label it is scored against—the resulting genome-wide rankings agree across the six tumour types at a Kendall concordance of 0.97. A gene’s transmissibility can therefore be anticipated before its protein is measured, which is what licenses extending the target map to cancer types with no tissue-referenced proteome of their own.

### A quantified funnel from transmissibility to accessible targets

With the transmission map and the predictor in hand, the full nomination path is a funnel, and every step is quantified from data (Figure 3). It begins with the 6,648 genes carrying both a measured and a predicted transmissibility across the CPTAC cohorts—so the target space is pan-cancer from the outset, not restricted to any one tumour type. Requiring transmission to protein (observed transmissibility ≥0.40) retains 3,104 genes. Independently, a per-(cohort, cytoband, gene) Fisher test identifies genes whose high tissue-relative protein is enriched in tumours where their band is amplified; 888 genes are co-elevated on a recurrent amplicon in at least one cohort (BH-FDR < 0.1), of which 541 are also transmitted. The last step keeps only genes an antibody could actually reach. An antibody–drug conjugate binds its target from outside the cell, so a useful target must be a membrane protein that displays a substantial portion of its structure—its ectodomain—on the cell surface, outside the lipid bilayer. We therefore apply a surface-accessibility gate: using UniProt topology annotation, we require at least 50 extracellular residues on a membrane-spanning protein. This gate yields the 22 nominated surface antigens, distributed across 18 distinct recurrent amplicons in 4 cancer types. The funnel is pan-cancer at every step; the antigens it yields are detailed next.

**Fig. 3.**
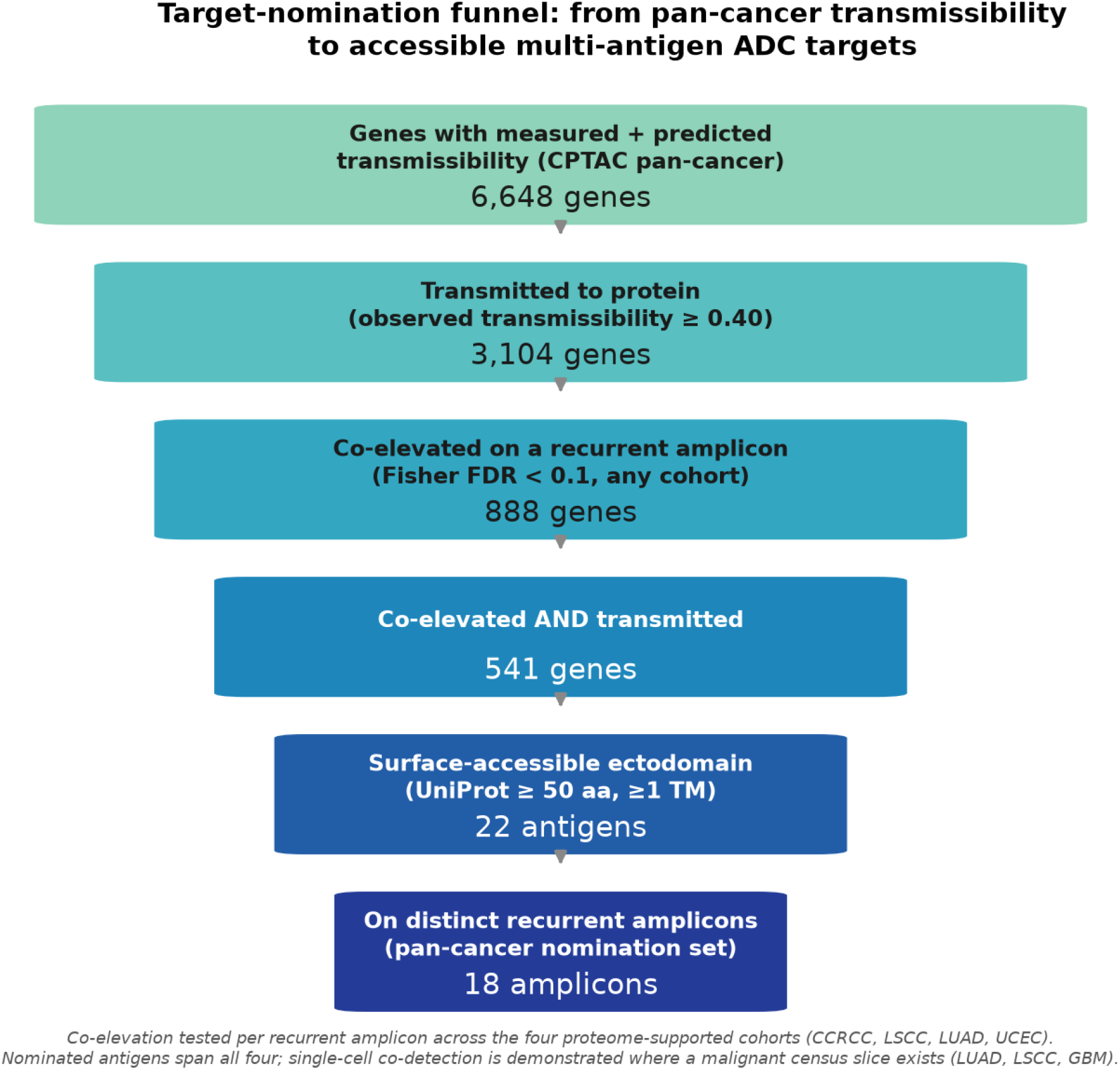
Target-nomination funnel. Each stage is computed: the transmissibility atlas, the per-cohort Fisher co-elevation test on recurrent cytoband amplicons (BH-FDR < 0.1), and the UniProt ectodomain gate; no counts are hardcoded. Co-elevation is evaluated in the four proteome-supported cohorts (CCRCC, LSCC, LUAD, UCEC) and the nominated antigens span all four.

### Nominated surface antigens and multi-antigen ADC constructs

A gene is a surface ADC target only if it is co-amplified on a recurrent amplicon, transmitted to protein, and presents an accessible extracellular ectodomain a native-format antibody could bind. Three “co-” terms operate at different biological levels and are kept distinct throughout: *co-amplified* means genes gained together in the same DNA segment; *co-elevated* means their proteins are jointly high across a cohort of tumours (a bulk, tumour-level test); and *co-detected* means both proteins are present in the same individual malignant cell (a single-cell test). We require all three of the nomination criteria: co-elevation on a recurrent amplicon (Fisher FDR < 0.1), measured transmissibility at or above 0.40, and a UniProt-annotated extracellular ectodomain of at least 50 residues on a membrane-anchored protein(19). Applied across every recurrent amplicon in the four proteome-supported cohorts, this nominates 22 surface antigens on 18 distinct amplicons spanning 4 cancer types (CCRCC, LSCC, LUAD, UCEC) (Table 1, Figure 4)—the nomination is pan-cancer, not restricted to lung. Examples span the amplicon landscape: EGFR, ITGB8 and TSPAN13 on lung 7p; NCSTN, HSD17B7 and MPZL1 on 1q (recurrent in LUAD, LSCC and UCEC); the transferrin receptor TFRC on the squamous 3q amplicon; the Wnt receptor FZD1 and integrin ITGB8 on clear-cell renal 7p/7q; and an NCSTN+MPZL1 set on endometrial 1q. The topology gate is decisive: it removes high-transmission genes with no accessible ectodomain (for example EFNA1, a GPI-anchored ligand), and downgrades multipass transporters even when amplified. Twenty-one of the 22 antigens are non-driver passengers; only EGFR (lung 7p) is a canonical oncogenic driver, and the passenger thesis does not depend on it (below). Two further non-driver antigens are broadly essential in the DepMap CRISPR screen (23Q4 Chronos gene-effect; TFRC, dependency -0.81; VMP1, -1.15). For an ADC this is not a liability on tumour—the payload kills the cell regardless of whether the antigen is essential—but essential, ubiquitously required genes are more likely to be expressed on normal tissue too, raising the on-target/off-tumour toxicity risk that the AND-gate and normal-tissue analysis below are designed to bound; we therefore flag these two rather than exclude them. The remaining 19 are neither drivers nor broadly essential. As orthogonal evidence for the surface call, 10 of the 22 — including the transmission leaders TFRC, NCSTN, MPZL1, ITGB8, PLXNA1 and TTYH3 — are listed on the experimental Cell Surface Protein Atlas(20) (mass-spectrometric surface capture); the remainder retain their UniProt topology call, with CSPA absence treated as weak evidence because surface capture sees only N-glycosylated proteins in the cell types assayed. Each antigen carries its driver, essentiality and CSPA flags in Table 1.

**Table 1.** Nominated surface antigens (pan-cancer) Antigen-level metrics (transmissibility, ectodomain, essentiality) are gene-intrinsic and pooled across the cohorts in which the gene is recurrently amplified, so a gene shared by several amplicons (e.g. NCSTN on 1q in LSCC, LUAD and UCEC) shows identical values on each row; Cohort gives the amplicon context, not a per-cohort remeasurement. Cohort = CPTAC cancer type in which the amplicon is recurrent; Measured / predicted = observed and gene-property-predicted transmissibility; n amp. = amplified cases underlying the measurement; Ecto = UniProt extracellular residues; TM = transmembrane helices; DepMap = mean CRISPR dependency effect (more negative = more essential); Class = driver / broadly essential (ess., DepMap ≤ -0.40) / passenger; CSPA = Cell Surface Protein Atlas category (high-confidence / putative / unspecific / — not listed).

| Cohort | Amplicon | Antigen | Meas. | Pred. | n amp. | Ecto | TM | DepMap | Class | CSPA |
| --- | --- | --- | --- | --- | --- | --- | --- | --- | --- | --- |
| CCRCC | CCRCC_5q | LNPEP | 0.67 | 0.49 | 54.0 | 894 | 1 | -0.03 | passenger | high |
| CCRCC | CCRCC_7p | ITGB8 | 0.65 | 0.32 | 195.0 | 642 | 1 | -0.08 | passenger | high |
| CCRCC | CCRCC_7q | FZD1 | 0.54 | 0.27 | 186.0 | 344 | 7 | 0.01 | passenger | — |
| LSCC | LSCC_12p | SLC2A3 | 0.59 | 0.34 | 90.0 | 82 | 12 | -0.02 | passenger | — |
| LSCC | LSCC_1q | NCSTN | 0.61 | 0.44 | 161.0 | 636 | 1 | 0.05 | passenger | high |
| LSCC | LSCC_1q | HSD17B7 | 0.53 | 0.30 | 156.0 | 229 | 1 | -0.13 | passenger | — |
| LSCC | LSCC_1q | MPZL1 | 0.51 | 0.38 | 156.0 | 127 | 1 | -0.16 | passenger | high |
| LSCC | LSCC_20q | TM9SF4 | 0.65 | 0.55 | 126.0 | 299 | 9 | -0.26 | passenger | — |
| LSCC | LSCC_3q | TFRC | 0.72 | 0.62 | 137.0 | 672 | 1 | -0.81 | ess. | high |
| LSCC | LSCC_3q | ATP11B | 0.66 | 0.47 | 148.0 | 80 | 10 | -0.13 | passenger | — |
| LSCC | LSCC_3q | PLXNA1 | 0.50 | 0.22 | 84.0 | 1218 | 1 | -0.08 | passenger | high |
| LSCC | LSCC_5p | LMBRD2 | 0.53 | 0.43 | 156.0 | 123 | 9 | 0.13 | passenger | put. |
| LSCC | LSCC_7p | TSPAN13 | 0.40 | 0.26 | 196.0 | 78 | 4 | 0.08 | passenger | put. |
| LSCC | LSCC_8p | SLC20A2 | 0.65 | 0.43 | 86.0 | 82 | 12 | -0.18 | passenger | — |
| LSCC | LSCC_8q | FZD6 | 0.50 | 0.39 | 130.0 | 274 | 7 | 0.14 | passenger | — |
| LUAD | LUAD_14q | TMX1 | 0.90 | 0.50 | 59.0 | 154 | 1 | -0.21 | passenger | — |
| LUAD | LUAD_16p | ABCC1 | 0.77 | 0.42 | 61.0 | 147 | 17 | -0.02 | passenger | high |
| LUAD | LUAD_17q | VMP1 | 0.85 | 0.55 | 80.0 | 166 | 7 | -1.15 | ess. | — |
| LUAD | LUAD_1q | NCSTN | 0.61 | 0.44 | 161.0 | 636 | 1 | 0.05 | passenger | high |
| LUAD | LUAD_5p | SLC12A7 | 0.83 | 0.43 | 160.0 | 181 | 12 | -0.02 | passenger | high |
| LUAD | LUAD_5p | CLPTM1L | 0.63 | 0.42 | 158.0 | 262 | 6 | 0.11 | passenger | put. |
| LUAD | LUAD_7p | TTYH3 | 0.66 | 0.42 | 193.0 | 275 | 5 | -0.05 | passenger | high |
| LUAD | LUAD_7p | ITGB8 | 0.65 | 0.32 | 195.0 | 642 | 1 | -0.08 | passenger | high |
| LUAD | LUAD_7p | EGFR | 0.59 | 0.36 | 202.0 | 621 | 1 | -0.35 | driver | high |
| LUAD | LUAD_7p | TSPAN13 | 0.40 | 0.26 | 196.0 | 78 | 4 | 0.08 | passenger | put. |
| UCEC | UCEC_1q | NCSTN | 0.61 | 0.44 | 161.0 | 636 | 1 | 0.05 | passenger | high |
| UCEC | UCEC_1q | MPZL1 | 0.51 | 0.38 | 156.0 | 127 | 1 | -0.16 | passenger | high |

**Fig. 4.**
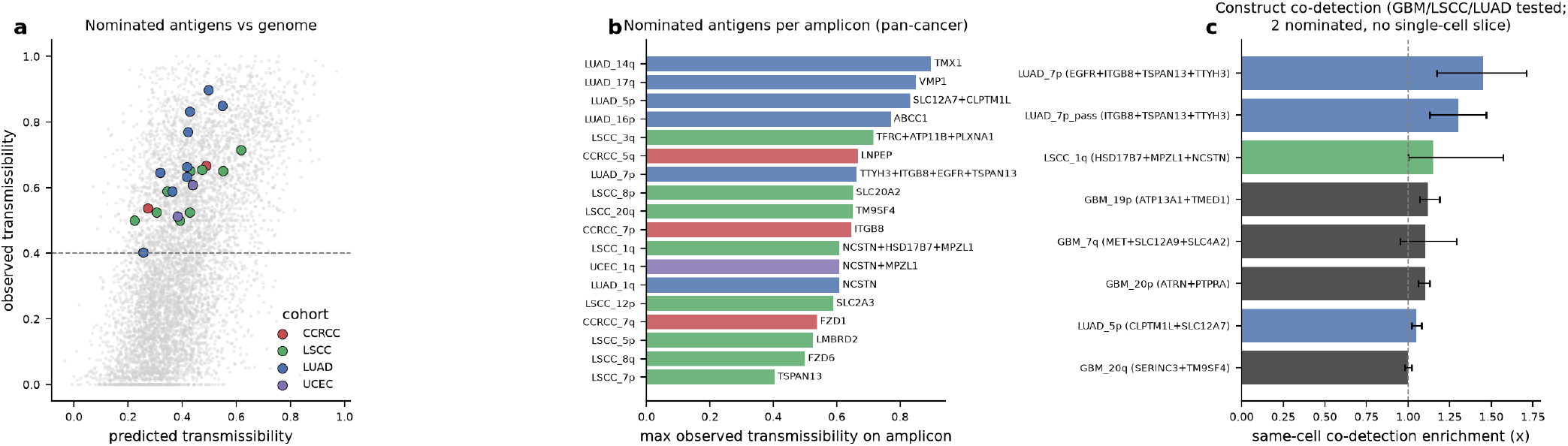
Pan-cancer surface targets. (a) The measured-nomination antigens (the four proteome-supported cohorts), coloured by cohort, placed against all 6,648 genes by predicted and observed transmissibility—they occupy the high-transmissibility regime above the 0.40 floor; glioblastoma is prediction-only and has no observed value, so it is not plotted here (its antigens are in Supplementary Table S4). (b) Nominated antigens per amplicon in those four cohorts, bar = maximum observed transmissibility on the amplicon. (c) Same-cell co-detection enrichment (depth-stratified null) for the constructs with a malignant single-cell slice (LUAD, LSCC measured; GBM prediction-only; 95% CI; significant constructs p < 0.001, GBM 20q n.s.); constructs on cohorts without a malignant single-cell atlas are nominated but not shown here.

Because the antigens on one amplicon rise together, an antibody that engages two or three of them can be built as an avidity AND-gate: a multispecific binder whose individual arms bind weakly, so that stable binding requires two or more antigens to be engaged at once, and a cell displaying only one is largely spared. We propose 10 multivalent constructs (Table 2), each assembled from the nominated antigens on a single amplicon (an amplicon with only one nominated antigen cannot form a construct); for LUAD 7p we additionally report the driver-excluded passenger-only set, which is the one carried into the experimental plan. Same-cell co-detection is measured directly in malignant single cells for the 8 constructs whose antigens are all present in the cancer type’s single-cell atlas slice (LUAD, LSCC and, for the prediction-only sets, GBM); the remaining 2 constructs (the LSCC 3q trio, whose PLXNA1 arm is absent from the LSCC slice, and the UCEC 1q pair, whose cohort has no malignant single-cell atlas) are nominated on the same genetic and topological evidence and flagged accordingly.

**Table 2.** Proposed multivalent ADC constructs. Same-cell enrichment is observed co-detection divided by a depth-matched expectation in malignant single cells (null permutes each antigen within per-cell sequencing-depth deciles; donor-block bootstrap 95% CI; depth-stratified permutation p). Evidence = measured (nominated from tissue-referenced proteomics) or prediction-only (nominated from predicted transmissibility because that cohort has no tissue-referenced proteome; GBM). Constructs on cohorts without a malignant single-cell atlas are nominated on genetic and topological evidence; co-detection cannot be evaluated there and they are labelled as such. The LUAD 7p amplicon is shown as two constructs: the full 4-valent set and the driver-excluded passenger-only trivalent set (ITGB8+TSPAN13+TTYH3), which is the therapeutic construct carried forward to the experimental plan, since the passenger thesis does not rely on the EGFR driver.

| Construct | Valence | Evidence | Same-cell enrichment (×) | 95% CI | perm p | Cells / donors | Single-cell tested |
| --- | --- | --- | --- | --- | --- | --- | --- |
| LUAD_7p (EGFR+ITGB8+TSPAN13+TTYH3) | 4-valent | measured | 1.45 | 1.17–1.71 | 0.001 | 41615 / 130 | yes |
| LUAD_7p passenger-only (ITGB8+TSPAN13+TTYH3) | trivalent | measured | 1.30 | 1.13–1.47 | 0.001 | 41615 / 130 | yes |
| LSCC_1q (HSD17B7+MPZL1+NCSTN) | trivalent | measured | 1.15 | 1.004–1.57 | 0.001 | 33234 / 42 | yes |
| GBM_19p (ATP13A1+TMED1) | bivalent | prediction-only | 1.12 | 1.07–1.19 | 0.001 | 390761 / 208 | yes |
| GBM_20p (ATRN+PTPRA) | bivalent | prediction-only | 1.1 | 1.06–1.13 | 0.001 | 390761 / 208 | yes |
| GBM_7q (MET+SLC12A9+SLC4A2) | trivalent | prediction-only | 1.1 | 0.95–1.29 | 0.001 | 390761 / 208 | yes |
| LUAD_5p (CLPTM1L+SLC12A7) | bivalent | measured | 1.05 | 1.02–1.08 | 0.001 | 41615 / 130 | yes |
| GBM_20q (SERINC3+TM9SF4) | bivalent | prediction-only | 1.0 | 0.98–1.02 | 0.3826 | 390761 / 208 | yes |
| LSCC_3q (ATP11B+PLXNA1+TFRC) | trivalent | measured | — | — | — | — | nominated |
| UCEC_1q (MPZL1+NCSTN) | bivalent | measured | — | — | — | — | nominated |

Two quantities matter here and should not be confused. The first is the *absolute* co-detection fraction—the share of malignant cells in which all of a construct’s antigens are simultaneously detected (by single-cell mRNA, a conservative proxy for surface protein; see Limitations)—which sets whether the construct engages a meaningful population of tumour cells. Because the individual antigens are detected in a large fraction of cells (typically 20–60% each), their joint co-detection is also substantial: 6.2% of LUAD cells co-detect all four LUAD 7p antigens and 17.2% co-detect the LUAD 5p pair (the LSCC 1q trio is lower at 1.2%). The second quantity is the *enrichment* over independence—how much co-detection exceeds the product of the single-antigen rates—which tests whether the antigens are coordinated rather than independently present. Enrichment is scored against a null that permutes each antigen within per-cell sequencing-depth deciles, so it reflects excess co-occurrence beyond what shared sequencing depth alone produces (a marginal-only null inflates it several-fold). On that stringent null the co-detected constructs are above independence with a bootstrap interval clear of 1.0 and permutation p < 0.001, at a modest 1.05–1.45×; one bivalent pair (GBM 20q) shows no enrichment. The two quantities carry different weight: the absolute fractions above show the constructs engage a real cell population, while the modest but significant enrichment shows that engagement is coordinated rather than coincidental. It is the direction and significance of the enrichment, not its absolute fold, that we treat as load-bearing.

### AND-gated co-targeting is selective in single cells

For a multivalent construct to be efficient on tumour, its antigens must appear together in the same malignant cell, not merely in the same tumour, and for it to be selective the co-occurrence must fall away in normal tissue. We test both in single malignant cells from three cancer types with a malignant single-cell atlas (41,615 LUAD cells from 130 donors, 33,234 LSCC cells from 42 donors, and 390,761 glioblastoma cells from 208 donors, from the CELLxGENE census(21)); the glioblastoma constructs are prediction-only (GBM has raw proteome in CPTAC but no tissue-referenced proteome, so it yields no measured surface nominations), and their single-cell test is a genuine out-of-sample check of the prediction pipeline. For each construct whose cancer type has a single-cell slice we ask whether its nominated antigens are co-detected in the same cell more often than independent detection would predict, scored against a depth-stratified null (each antigen permuted within per-cell sequencing-depth deciles, so the null preserves the depth structure that otherwise inflates co-detection). Of the 8 tested constructs, 6 are co-detected above independence (1.05–1.45×), each with permutation p < 0.001 and a donor-block bootstrap interval clear of 1.0; 1 more (the trivalent GBM 7q set) is permutation-significant but with a wide bootstrap interval that spans 1.0 (0.95–1.29), so its enrichment is not established, and 1 (the bivalent GBM 20q pair) is not significant on this null (1.00×, p = 0.38). The magnitudes are modest — a marginal-only null returns several-fold higher values, but most of that is the depth artefact the stratified null removes — and the higher-valence LUAD 7p (four antigens) set retains the largest true excess (1.45×), consistent with amplicon-driven coordination. Critically, LUAD 7p survives removal of its one driver, EGFR: the passenger-only ITGB8+TSPAN13+TTYH3 set is still co-detected at 1.30× (95% CI 1.13–1.47, p < 0.001), so the co-targeting rests on passengers, not on the driver. The glioblastoma constructs — nominated without any GBM surface-abundance measurement, from predicted transmissibility alone — reproduce the same pattern: 2 of the 4 GBM constructs (19p ATP13A1+TMED1 at 1.12×, 20p ATRN+PTPRA at 1.10×) are co-detected above independence with intervals clear of 1.0, so the full predict→surface-gate→single-cell-verify path closes in a cancer type whose proteome is not tissue-referenced and so drives no measured nomination. We stress that prediction-only nomination is a lower-sensitivity filter than measurement: because the predictor explains about a third of per-gene variance, applying it alone recovers the coordinated, high-transmissibility genes but will miss genuine targets that measurement would have caught—indeed several antigens nominated in the proteome-supported cohorts (for example ITGB8, TSPAN13) fall below the predicted-transmissibility threshold and would not have been nominated from prediction alone. The GBM result therefore demonstrates that the pipeline *can* close end-to-end without proteomics, not that prediction matches measurement. The load-bearing result is the direction and significance of same-cell co-occurrence, not any single fold value.

The off-tumour side is where AND-gating earns its selectivity. A normal cell binds an avidity AND-gate only if its weakest (limiting) antigen clears the binding threshold. Raising the per-antigen threshold from detection to binding-relevant levels collapses the fraction of normal cell types in which all of a construct’s antigens are co-present toward zero, provided each construct contains at least one antigen that is low in normal tissue(22, 23). Together these give the selectivity argument its two halves: measurable on-tumour same-cell co-occurrence, and threshold-gated off-tumour sparing (Figure 5).

**Fig. 5.**
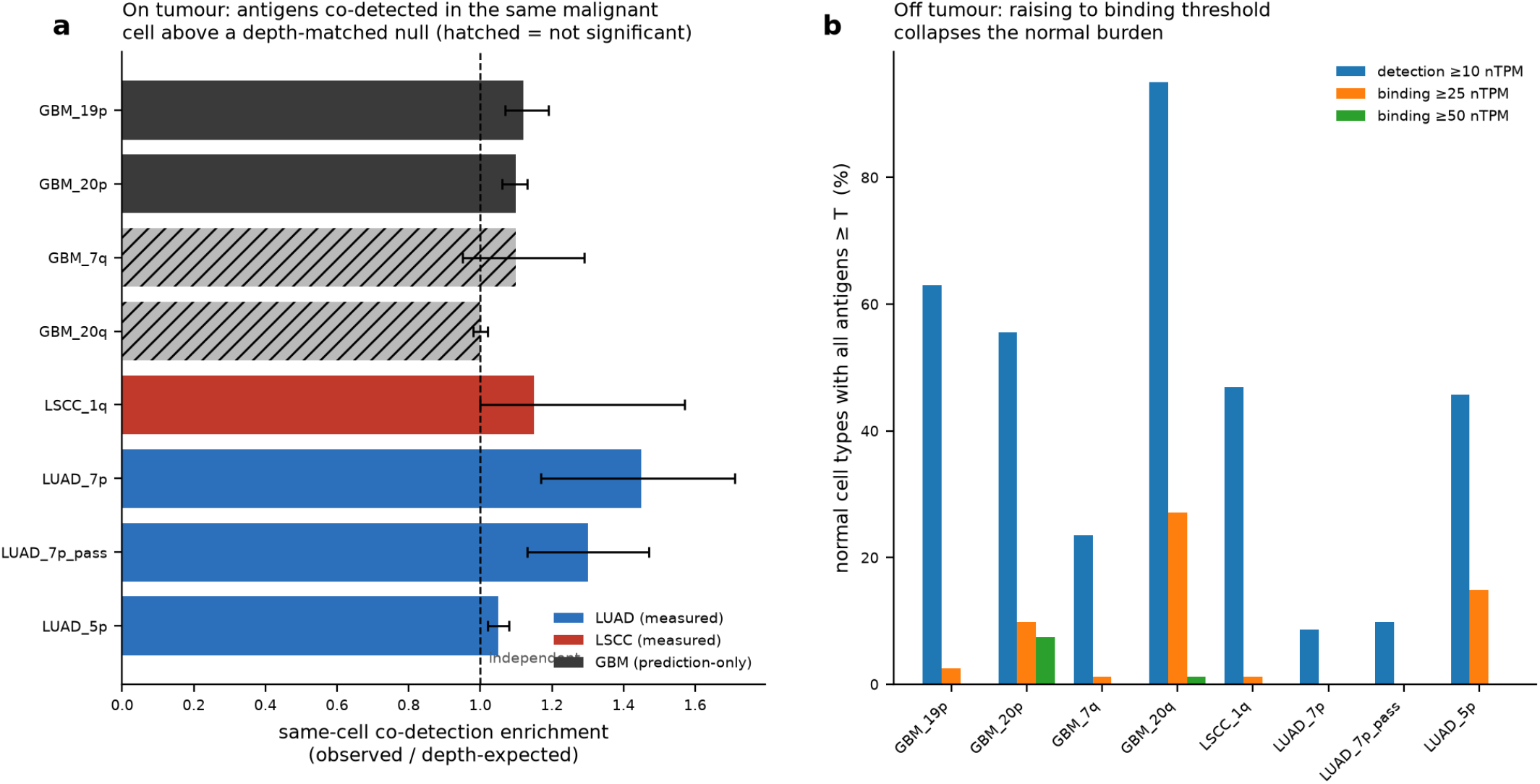
AND-gated selectivity in single cells, for the nominated constructs whose cohort has a malignant single-cell slice (LUAD: 41,615 cells / 130 donors; LSCC: 33,234 cells / 42 donors; GBM: 390,761 cells / 208 donors; CELLxGENE). (a) Same-cell co-detection enrichment in malignant cells for each construct (observed / depth-expected co-detection; donor-block bootstrap 95% CI, 2,000 resamples; depth-stratified permutation p, 1,000 permutations, each antigen shuffled within per-cell sequencing-depth deciles). Constructs are shown for the three cohorts with a malignant single-cell atlas (LUAD, LSCC measured; GBM prediction-only); those above independence span 1.05–1.45× (p < 0.001, interval clear of 1.0), while the GBM 7q trio has a wide interval spanning 1.0 and the GBM 20q pair is not significant. A marginal-only null returns several-fold higher values, most of which is the depth artefact this null removes (see text). (b) Fraction of Human Protein Atlas normal cell types in which all antigens of a construct exceed a per-antigen threshold, at detection (10 nTPM) versus binding-relevant (25, 50 nTPM) levels; raising the threshold collapses the normal-tissue burden (LUAD 7p to zero), which is the basis of the AND-gate selectivity window.

### A plan to test the nominated sets

The nominations are hypotheses about surface protein, and the framework makes each one falsifiable. The forward plan is a staged go/no-go (Figure 6): confirm surface co-expression of a set’s antigens in amplicon-positive versus amplicon-negative cell lines; measure native-surface antibody binding for marginal ectodomains; build an avidity AND-gate construct and confirm that tumour killing requires both antigens while single-antigen cells are spared; and measure the selectivity window against matched normal tissue. A no-go at any step returns the set to nomination with the failing antigen dropped, which the confidence tiering makes inexpensive. Concrete entry points are amplicon-positive cell lines identified from DepMap 23Q4 copy number (all anchor genes at relative copy number ≥1.4 in the matching lineage): the LUAD 7p set (ITGB8+TSPAN13+TTYH3, driver EGFR excluded) in NCI-H3255, NCI-H1838 and HCC4006, with the amplicon-negative LUAD lines NCI-H441 and NCI-H358 as single-antigen controls; the LSCC 1q set (HSD17B7+MPZL1+NCSTN) in LUDLU-1/GT3TKB; and the squamous 3q set (ATP11B+PLXNA1+TFRC) in the 3q-amplified lines HCC95 and LC-1/sq. The LUAD 7p and LSCC 1q entry points additionally have their same-cell co-detection confirmed here on the depth-stratified null (Figure 5).

**Fig. 6.**
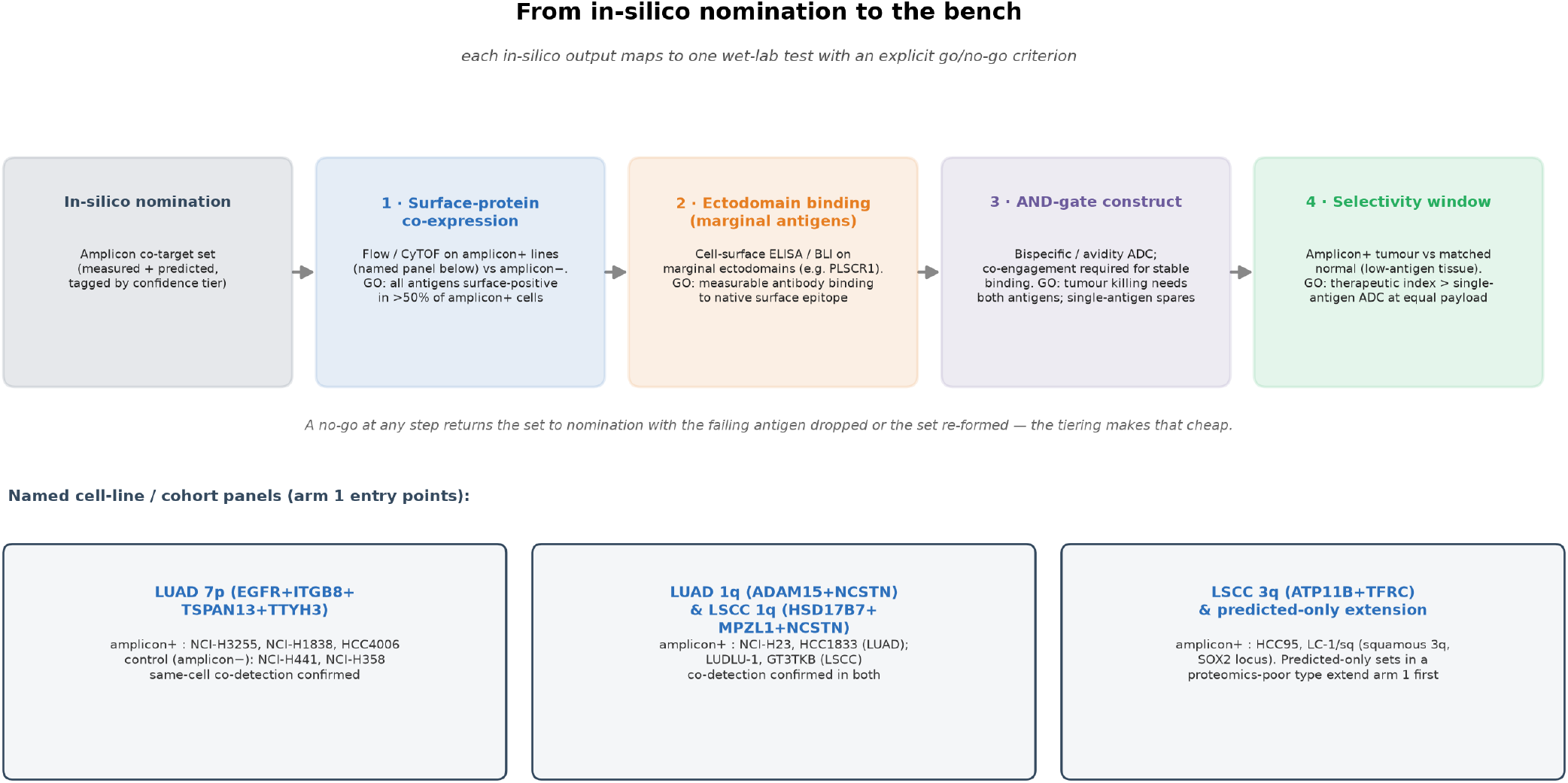
From in-silico nomination to the bench. Each in-silico output maps to one wet-lab test with an explicit go/no-go criterion, with named cell-line and cohort entry points.

## Discussion

We have built an integrated, quantified map of how a copy-number amplification reaches the cell surface, and used it to nominate multi-antigen ADC co-target sets. Three features make it more than a target list. First, it is a full-path account: the framework resolves the amplicon-to-mRNA-to-protein path into its steps and shows where the between-gene ranking is set—gated at chromatin and transcription, where, among the regulatory covariates we measure directly, promoter methylation and accessibility each explain an order of magnitude more variance than gene structure, and then attenuated post-transcriptionally (Figure 1)—so the map addresses not just which genes transmit but where the control lies. Second, transmissibility is intrinsic and therefore portable—predictable from gene biology alone, not positional, and stable across lineages—so the map extends by prediction to cancer types that lack a tissue-referenced proteome. Where both a measurement and a prediction exist, an empirical-Bayes posterior (Supplementary Methods) makes explicit how much of each per-gene estimate is measured and how much predicted; it is a refinement for thin-proteomics cohorts and does not gate the nominations, which rest on observed transmissibility and the co-elevation test directly. Third, the modality and the genetics fit: co-amplified passengers rise together with an amplicon the tumour is selected to keep, and avidity AND-gating turns that co-occurrence into selectivity, which we confirm at single-cell resolution.

This has a direct consequence for how widely a nominated set should apply within a cancer type. Once an amplicon is present, which of its genes reach the surface as protein is set by the regulatory gates quantified in Figure 1—promoter DNA methylation and chromatin accessibility—and both of these are predominantly properties of the cell of origin rather than of the individual tumour: across the normal human cell types profiled in a reference methylation atlas, biological replicates of the same cell type are more than 99% identical in methylation(24), and primary-tumour chromatin accessibility clusters predominantly by lineage(25). Because we deliberately nominate low-essentiality passenger antigens with no role in the cancer’s biology, there is little tumour-specific selection acting on their expression to override those shared gates. Two tumours of the same type that both carry the same amplicon therefore start from the same cell-of-origin gate settings and face no divergent selective pressure on the passengers, so they should present substantially the same set of amplicon-driven surface antigens. Our transmissibility values already quantify this recurrence directly: transmissibility is the fraction of amplified tumours in a cohort in which a gene’s protein is elevated, so a nominated antigen with transmissibility 0.6–0.9 is, by definition, one whose amplification reaches the surface in a high proportion of patients carrying that gain in that cancer type. The practical implication is that a construct nominated against, say, the lung-squamous 3q amplicon is expected to be actionable across a large share of 3q-gained lung-squamous patients, not only the tumour in which it was measured. This is a within-cancer-type, across-patient claim; it is distinct from and does not depend on the weaker cross-lineage transfer of the transmissibility ranking. The measured cross-cohort correlations bear this out: observed transmissibility correlates most strongly between the two lung subtypes (Spearman *ρ* = 0.88 over 3,653 shared genes) and remains positive and substantial across every one of the 15 cohort pairs (all *ρ* = 0.60–0.88, each p ≪ 10^*−*30^), with the highest values among the more closely related epithelial lineages — the gradient expected if the gates are set largely by cell of origin. It is this within- and near-lineage portability, not a universal cross-cancer constant, that additionally licenses extending the map by prediction to related cancer types with no tissue-referenced proteome of their own.

The clinical reading is deliberately measured. The nominations are transcript- and prediction-level calls about surface protein, not surface-protein measurements; the honest bound is that gene properties explain about a third of per-gene transmission variance, so the framework ranks the genome well but will mispredict individual genes. That is why every lead enters a staged wet-lab test before it could be called a target, and why the confidence tiers—measured, predicted, or both—travel with every candidate rather than being averaged away.

## Data Availability

All data produced are available online at https://github.com/ucl-respiratory/amplicon-targeting

## Limitations

Several bounds are explicit. The transmissibility prior explains about a third of per-gene variance; it is a genome-wide prior, not a per-gene oracle, and individual genes are mis-predicted. The surface calls rest on UniProt topology annotation, not on measured surface abundance, and the next step for any lead is direct surface-protein and tumour-versus-normal measurement. The single-cell co-detection uses detection (non-zero counts), which is conservative for abundance but does not by itself establish that both antigens reach binding-relevant surface density; the normal-tissue threshold analysis addresses magnitude on the off-tumour side but the on-tumour density claim still requires protein measurement. Co-detection is scored against a null that controls for per-cell sequencing depth, which reduces the enrichments to modest excess (1.05–1.45×) and renders one bivalent pair non-significant; the argument rests on the direction and significance of same-cell co-occurrence together with the threshold-gated normal sparing, not on large fold values. The nomination is also amplification-threshold dependent: at a stringent high-level cut (adjusted copy number ≥2.0) the recurrent-amplicon set and the co-elevated-and-transmitted set shrink substantially and only 6 of the 22 nominated antigens survive (Supplementary Table S3); we use ≥1.4 because the clinical premise is the broad, near-ubiquitous segmental gains that define these amplicons, not rare focal high-level events. The chromatin gate is quantified from both promoter DNA methylation (GDC EPIC arrays, mapped to genes through the Illumina manifest) and tumour ATAC accessibility, which prove complementary.

## ACKNOWLEDGEMENTS

This work was undertaken as part of the Claude Science hackathon in July 2026, sponsored by Anthropic and Gladstone Institutes. We are grateful for their support. The funders had no role in the design of the study, the analysis or interpretation of the results, or the decision to publish.

## Supplementary Methods

### Data and cohorts

Matched copy number, mRNA and protein were assembled across six tumour types (LSCC, LUAD, GBM, PDA, CCRCC, UCEC) from public proteogenomic resources(14, 15), harmonised to a common gene- and case-level table. Amplification is defined on a ploidy-adjusted copy-number basis (adjusted copy number ≥1.4), the regime relevant to the clinical output. Tumour chromatin accessibility is taken from cancer-type-matched ATAC-seq(25); single-cell malignant-cell profiles are from the CELLxGENE census(21) (lung adenocarcinoma, lung squamous carcinoma and glioblastoma, primary malignant cells); normal single-cell and bulk expression are from the Human Protein Atlas(22, 23) and GTEx(26). Surface topology is from UniProt reviewed human entries(19) (extracellular topological-domain residues and transmembrane-helix count). The full index of datasets and how each is used is in Supplementary Table S2.

### Transmission cascade

Per gene and per tumour type, copy-number-to-mRNA transmission and copy-number-to-protein responsiveness are Pearson correlations on the joint non-missing cases (minimum 20 cases per type, at least two types, Fisher-z averaging weighted by n-3); attenuation is their difference. The chromatin gate is a nested cross-validated linear model of pergene transmission, comparing gene structure alone against structure plus promoter DNA methylation, plus tumour promoter accessibility, and plus both, across the profiled cancer types. Promoter methylation is the mean SeSAMe beta over a gene’s TSS200/TSS1500 probes on the Illumina EPIC array (GDC CPTAC-3, 655 matched tumours); probes are mapped to genes through the Illumina EPIC manifest (UCSC_RefGene annotation) rather than through ChAMP, which does not build on current R/Bioconductor. Accessibility is the tumour-type promoter ATAC signal (TCGA-ATAC).

### Transmissibility predictor

A gradient-boosted regressor(27) (600 trees, depth 4, learning rate 0.03, subsample and column-sample 0.8, seed 2) predicts per-gene transmissibility from the gene-property features defined in Supplementary Table S1, with no protein-derived input. Generalisation is estimated by five-fold leave-gene-out out-of-fold prediction; the positional control refits holding out whole chromosome arms in turn; cross-lineage transfer is the Kendall concordance of predicted rankings across leave-one-lineage-out refits. No protein-derived feature is used as a predictor.

### Empirical-Bayes combination of measurement and prediction

Direct co-elevation is precise where proteomics is deep and noisy where it is thin, while the predictor is available everywhere but explains about a third of per-gene variance. We combine them in an empirical-Bayes posterior(28) that shrinks each per-gene measurement toward the predictor prior in inverse proportion to the measurement’s precision. The posterior for each gene is a precision-weighted blend of the predictor prior (mean) and the measured transmissibility, with binomial measurement variance p(1-p)/n set by the number of amplified cases and a between-gene prior variance estimated by method of moments. The thin-cohort recovery test treats genes with at least 150 amplified cases as reference truth (n = 6,648 genes total), injects binomial noise at a grid of cohort sizes, and scores raw measurement, prior-only and posterior against the reference by rank correlation and RMSE over repeated resamples (seed 2). The posterior never falls below either the raw measurement or the prior at any cohort size; the raw measurement overtakes the prior-only floor only above about 5 amplified cases; and the prior-only floor (*ρ* = 0.49) is the achievable ceiling when no proteomics exists at all (Supplementary Figure S1). This yields a posterior transmissibility for every gene with an explicit decomposition of how much is measured versus predicted; the nomination funnel itself uses observed transmissibility and the co-elevation test directly, so the posterior is a refinement for thin-proteomics cohorts rather than a gate on the headline results.

### Recurrent amplicons, co-elevation and surface nomination

A gene is amplified in a tumour when its ploidy-adjusted copy number is at least 1.4 (deduplicated to one value per case). A cytoband is amplified in a tumour when at least 50% of its genes are amplified, and recurrent in a cohort when it is amplified in at least 20% of tumours (minimum 8). A gene is co-elevated on a recurrent amplicon when its tissue-referenced protein rank exceeds 0.80 (“high”) in amplified cases significantly more often than in non-amplified cases, by a one-sided Fisher exact test; p-values are Benjamini–Hochberg-corrected within cohort and controlled at FDR < 0.10. This band-level test is run in the four cohorts with tissue-referenced proteome (CCRCC, LSCC, LUAD, UCEC; GBM has copy number but no tissue-referenced protein and PDA has no cytoband meeting the recurrence bar, so neither enters the co-elevation test). Nominated antigens are co-elevated genes (FDR < 0.10) with observed transmissibility ≥0.40 that pass a live UniProt topology gate—at least 20 extracellular residues on a membrane-anchored protein to be scored accessible, with ≥50 required for nomination as a confidently accessible epitope. Because transmissibility and co-elevation are both derived from the same tissue-referenced protein-abundance indicator (protein rank > 0.80), they are not independent filters but two views of the same protein signal — transmissibility summarising its level across amplified tumours, co-elevation testing its enrichment in amplified versus non-amplified cases — so the surface-topology gate is the one orthogonal criterion; the experimental Cell Surface Protein Atlas(20) is used as a corroborating flag, not a gate. Each antigen is tagged by measured and predicted evidence, driver and essentiality status, CSPA membership, and its DepMap dependency effect. Same-cell co-detection enrichment is the observed fraction of malignant cells co-detecting all antigens of a set divided by the product of per-antigen detection rates, with donor-block bootstrap 95% intervals (2,000 resamples) and a depth-stratified permutation p (1,000 permutations; each antigen column shuffled independently within per-cell sequencing-depth deciles, so the null preserves both per-gene detection rate and per-cell depth and the enrichment measures co-detection beyond what shared depth alone produces), computed in the LUAD, LSCC and GBM malignant single-cell slices (CELLxGENE: 41,615 cells / 130 donors, 33,234 cells / 42 donors and 390,761 cells / 208 donors; GBM is prediction-only and its single-cell test is an out-of-sample check of the prediction pipeline). Because a higher-valence set has a smaller independence baseline, the enrichment ratio scales with the number of antigens; the load-bearing claim is the direction and significance of co-detection, not the absolute fold. Normal-tissue burden is the fraction of Human Protein Atlas normal cell types in which the limiting antigen of a set clears a per-antigen threshold, evaluated at detection (10 nTPM) and binding-relevant (25, 50 nTPM) levels.

**Fig. S1.**
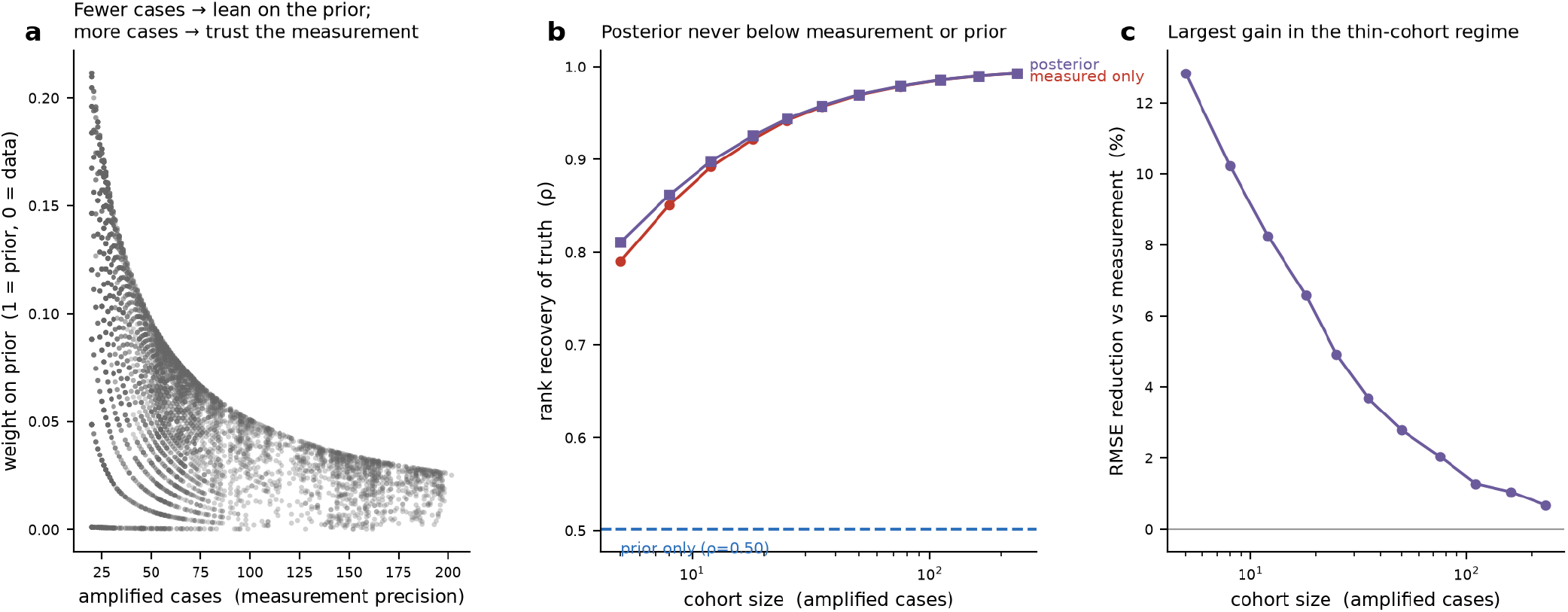
Empirical-Bayes combination of measured and predicted transmissibility (n = 6,648 genes). (a) Weight placed on the prior falls as the number of amplified cases (measurement precision) rises. (b) Thin-cohort recovery of reference truth (genes with ≥ 150 amplified cases): the posterior tracks or beats the raw measurement and never drops below the prior-only floor (*ρ* = 0.49). (c) RMSE reduction of the posterior over raw measurement, largest in the thin-cohort regime.

### Code availability

All code is available at https://github.com/ucl-respiratory/amplicon-targeting. A data-preparation pipeline fetches every input from its primary public source, and an analysis pipeline computes every figure, table and in-text value from those cached layers.

### Reproducibility

All analysis code is in the integrated pipeline folder, with a single configuration module fixing thresholds and seeds (seed 2 throughout), a data preparation driver that caches the source layers, and one script per figure. Every figure regenerates deterministically from the cached source layers.

Every feature used by the transmissibility predictor, grouped by family. No feature is derived from protein measurement; only the predicted quantity (transmissibility) is protein-derived. Column names match config.py (PREDICTOR_GROUPS) and the exported feature table.

Every external dataset the pipeline consumes, its source, size, and role in the analysis.

The full nomination funnel recomputed at the working amplification threshold (adjusted copy number ≥1.4, the broad-gain regime the clinical premise rests on) and at a stringent high-level threshold ( ≥2.0). High-level focal amplification is rarer, so every funnel stage contracts and only 6 of the 22 nominated antigens survive at ≥2.0; the broad-gain threshold is used because near-ubiquitous segmental gains, not rare focal events, are the actionable target.

Glioblastoma has copy number, mRNA and raw proteome in CPTAC — and so contributes to the transmission cascade — but its proteome is not referenced to a matched normal tissue of origin, the quantity co-elevation requires, so GBM contributes no measured surface nomination. Running the gene-property predictor end-to-end on GBM copy number alone (recurrent amplicon → predicted transmissibility ≥0.40 → UniProt surface gate) nominates the antigens below without using any GBM protein measurement. As a face-validity check, the two canonical GBM receptor-tyrosine-kinase amplicon oncogenes, EGFR (7p) and MET (7q), are among the genes recovered—reassuring, though both sit close to the nomination threshold and are the most obvious genes on their amplicons, so their recovery is a sanity check rather than strong validation. Of the 11 nominations, 5 are confirmed on the experimental Cell Surface Protein Atlas. Assembled into multivalent constructs and tested in 390,761 malignant GBM cells (208 donors), the 19p (ATP13A1+TMED1) and 20p (ATRN+PTPRA) sets are co-detected above independence on the depth-stratified null (1.12× and 1.10×, p < 0.001, intervals clear of 1.0) and the 7q (MET+SLC12A9+SLC4A2) set is permutation-significant — so the full predict → surface-gate → single-cell-verify path closes without any GBM surface-abundance measurement: the nomination uses gene-intrinsic prediction, the surface call uses UniProt topology, and the verification uses single-cell detection — none of them a GBM tissue-referenced proteome.

Observed (measured, not predicted) transmissibility computed independently in each proteome-supported cohort, then correlated between cohort pairs (Spearman, genes with 20 amplified cases in both). Transfer is strong within lineage (the two lung subtypes), moderate to related epithelium, and weak to the distant clear-cell renal cohort — the gradient expected if the regulatory gates are set by cell of origin. This is the empirical basis for the within-cancer-type portability claim in the Discussion.

**Table S2.**
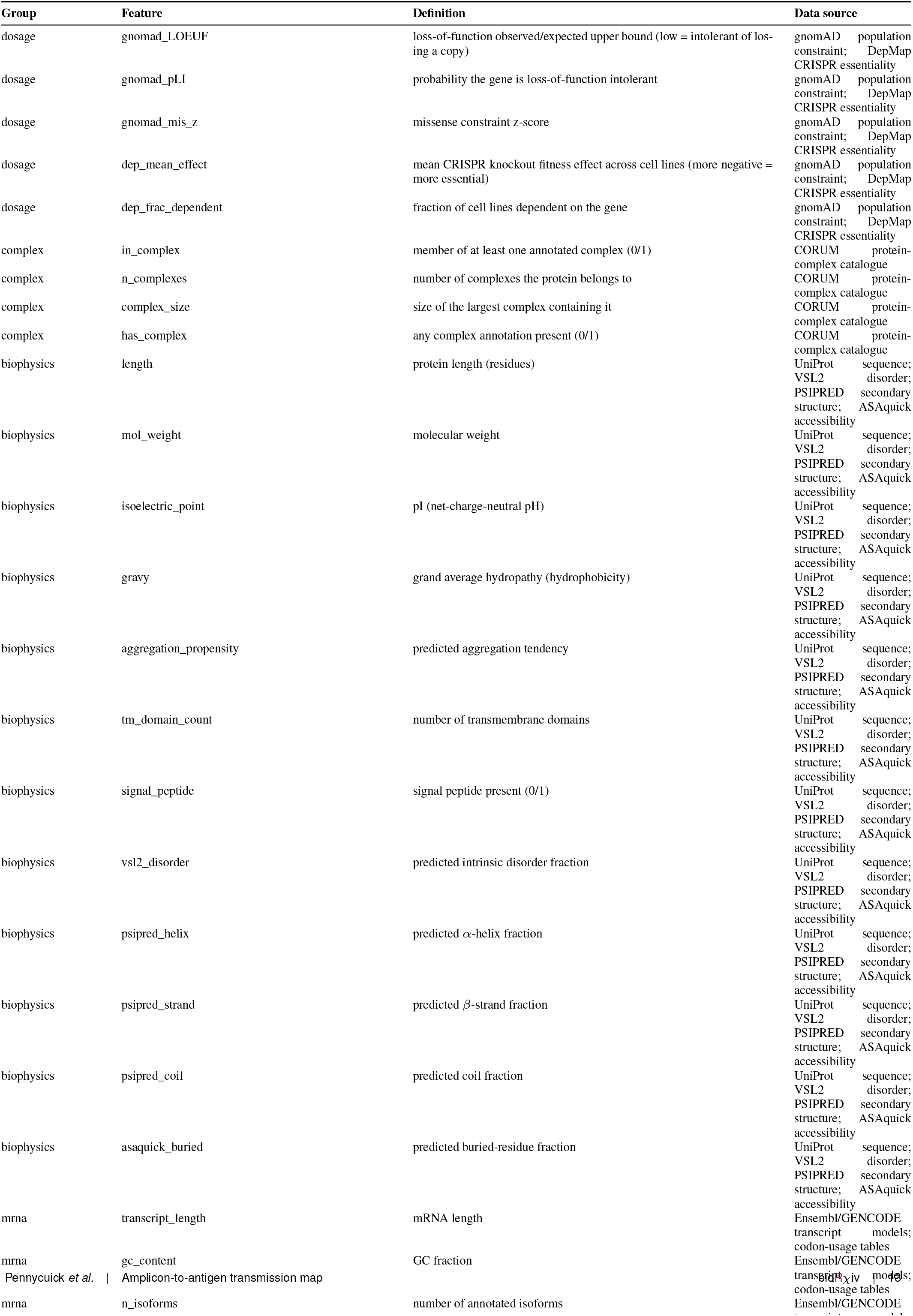
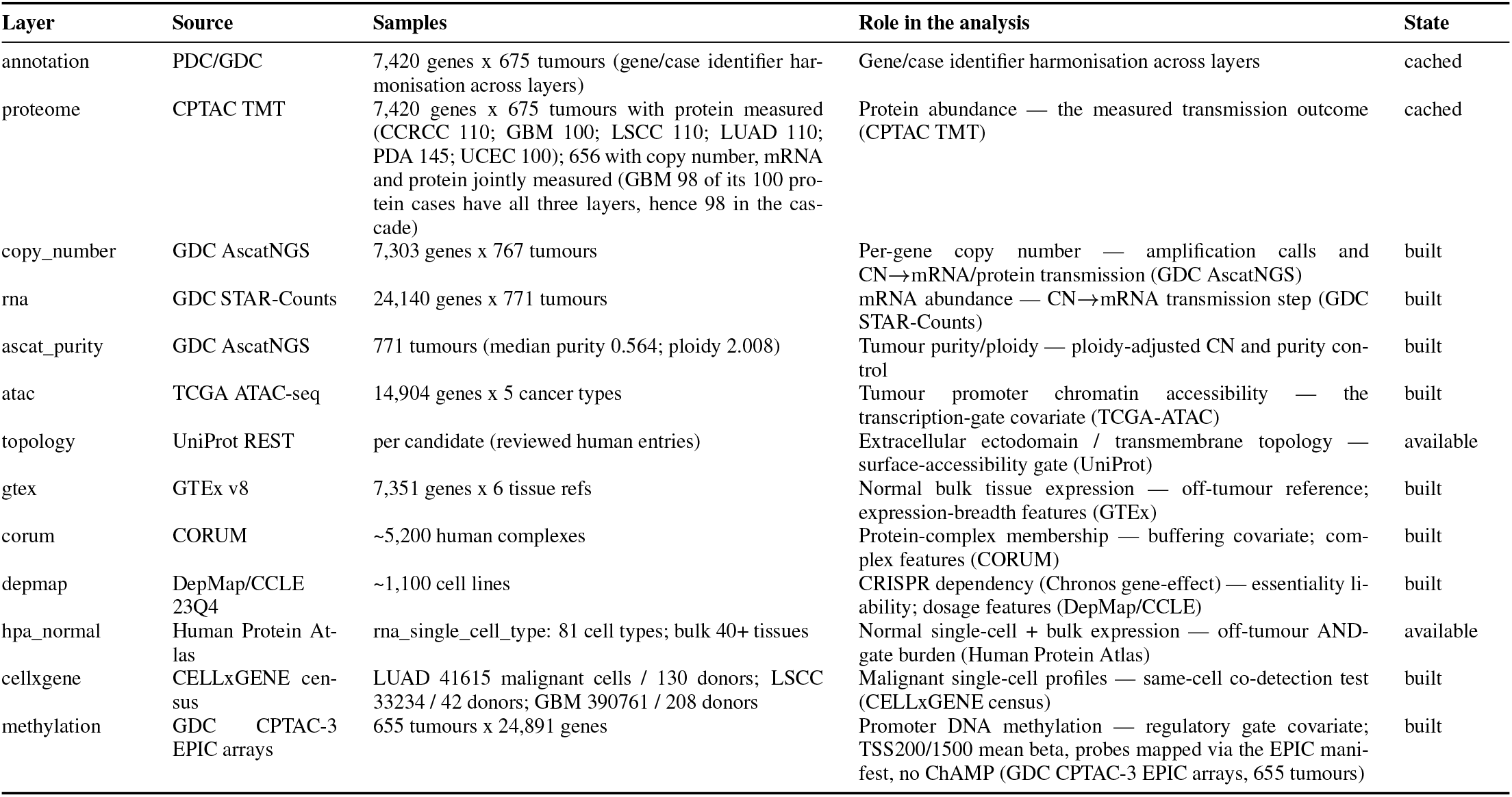
Index of datasets and how each is used. Sample counts are the exact parsed dimensions for each layer; counts marked ‘∼’ are rounded totals for whole reference databases (CORUM complexes, DepMap cell lines).

**Table S3.**
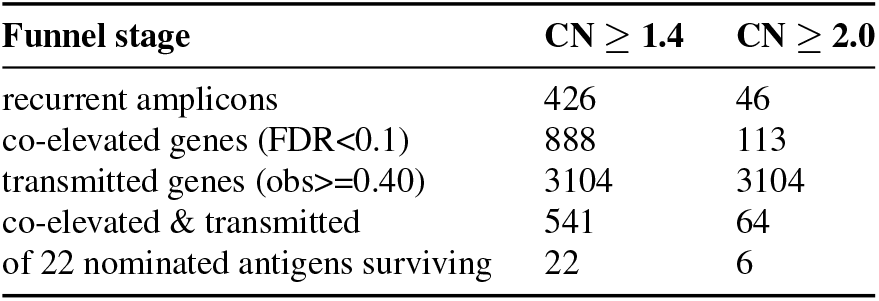
Amplification-threshold sensitivity. Funnel stage counts recomputed end-to-end from source at two copy-number thresholds for calling a gene amplified (cnadjusted ≥ 1.4, the paper basis, and ≥ 2.0, a stringent alternative). The recurrent-amplicon, co-elevation and co-elevated-and-transmitted stages tighten sharply at 2.0; the transmitted-gene count is identical at both because transmissibility is defined on a fixed high-level amplification filter (copy number ≥ round(ploidy)+1 and ≥ 3), independent of this amplicon-calling threshold. The paper uses ≥ 1.4 throughout; the nomination survives at 2.0 for a reduced set of antigens.

**Table S4.** Prediction-only nominations in a cohort with no tissue-referenced proteome (GBM)

| Antigen | Arm | GBM amp. freq. | Pred. transmiss. | Ecto (aa) | TM | CSPA |
| --- | --- | --- | --- | --- | --- | --- |
| TMED1 | 19p | 0.23 | 0.57 | 171 | 1 | — |
| ATP13A1 | 19p | 0.27 | 0.41 | 225 | 10 | — |
| LSR | 19q | 0.26 | 0.41 | 259 | 1 | — |
| ATRN | 20p | 0.30 | 0.47 | 1196 | 1 | yes |
| PTPRA | 20p | 0.30 | 0.46 | 132 | 1 | yes |
| TM9SF4 | 20q | 0.28 | 0.46 | 299 | 9 | — |
| SERINC3 | 20q | 0.27 | 0.45 | 179 | 8 | — |
| EGFR | 7p | 0.76 | 0.42 | 621 | 1 | yes |
| SLC12A9 | 7q | 0.68 | 0.47 | 368 | 12 | yes |
| MET | 7q | 0.71 | 0.45 | 908 | 1 | yes |
| SLC4A2 | 7q | 0.70 | 0.44 | 53 | 10 | — |

**Table S5.** Observed cross-cohort transmissibility transfer.

| Cohort pair | Shared genes | Spearman $\rho$ | p |
| --- | --- | --- | --- |
| LSCC-LUAD | 3653 | 0.88 | 0 |
| LUAD-UCEC | 1507 | 0.81 | 0 |
| LSCC-UCEC | 1514 | 0.79 | 0 |
| GBM-LUAD | 867 | 0.73 | 1.7e-142 |
| GBM-LSCC | 1178 | 0.71 | 2.2e-178 |
| CCRCC-LSCC | 1257 | 0.68 | 1.7e-172 |
| CCRCC-GBM | 452 | 0.68 | 3.1e-62 |
| PDA-UCEC | 639 | 0.67 | 1.5e-85 |
| LSCC-PDA | 964 | 0.67 | 7.7e-128 |
| CCRCC-LUAD | 1748 | 0.67 | 1.4e-225 |
| LUAD-PDA | 910 | 0.66 | 1.0e-115 |
| GBM-UCEC | 691 | 0.64 | 1.1e-80 |
| CCRCC-PDA | 326 | 0.63 | 2.8e-37 |
| GBM-PDA | 415 | 0.61 | 4.9e-44 |
| CCRCC-UCEC | 690 | 0.60 | 2.7e-68 |

## Bibliography

1. Nelson BE, et al. Leveraging TROP2 antibody-drug conjugates in solid tumors. Annu Rev Med. 2024;75:31–46. doi:10.1146/annurev-med-071322-065903

2. Gazzah A, et al. Safety, pharmacokinetics, and antitumor activity of the anti-CEACAM5-DM4 antibody-drug conjugate tusamitamab ravtansine (SAR408701) in patients with advanced solid tumors: first-in-human dose-escalation study. Ann Oncol. 2022;33:416–25. doi:10.1016/j.annonc.2021.12.012

3. Izalontamab brengitecan, an EGFR and HER3 bispecific antibody-drug conjugate, versus chemotherapy in heavily pretreated recurrent or metastatic nasopharyngeal carcinoma: a multicentre, randomised, open-label, phase 3 study. Lancet. 2025. doi:10.1016/S0140-6736(25)01954-3

4. McCaughan F, et al. Progressive 3q amplification consistently targets SOX2 in preinvasive squamous lung cancer. Am J Respir Crit Care Med. 2010;182:83–91. doi:10.1164/rccm.201001-0005OC

5. Jeon S, et al. Chromosome 3q amplification in lung squamous cell carcinoma. Thorac Cancer. 2023;14:2325–34. doi:10.1111/1759-7714.15045

6. Gonçalves E, et al. Widespread post-transcriptional attenuation of genomic copy-number variation in cancer. Cell Syst. 2017;5:386–98. doi:10.1016/j.cels.2017.08.013

7. Vogel C, Marcotte EM. Insights into the regulation of protein abundance from proteomic and transcriptomic analyses. Nat Rev Genet. 2012;13:227–32. doi:10.1038/nrg3185

8. Stingele S, et al. Global analysis of genome, transcriptome and proteome reveals the response to aneuploidy in human cells. Mol Syst Biol. 2012;8:608. doi:10.1038/msb.2012.40

9. Sousa A, et al. Multi-omics characterization of interaction-mediated control of human protein abundance levels. Mol Cell Proteomics. 2019;18:S114–25. doi:10.1074/mcp.RA118.001280

10. Protein buffering of aneuploidy is driven by coordinated factors identified through machine learning. Mol Syst Biol. 2026. doi:10.1038/s44320-026-00187-9

11. Beroukhim R, et al. The landscape of somatic copy-number alteration across human cancers. Nature. 2010;463:899–905. doi:10.1038/nature08822

12. Zack TI, et al. Pan-cancer patterns of somatic copy number alteration. Nat Genet. 2013;45:1134–40. doi:10.1038/ng.2760

13. Sanchez-Vega F, et al. Oncogenic signaling pathways in The Cancer Genome Atlas. Cell. 2018;173:321–37. doi:10.1016/j.cell.2018.03.035

14. Gillette MA, et al. Proteogenomic characterization reveals therapeutic vulnerabilities in lung adenocarcinoma. Cell. 2020;182:200–25. doi:10.1016/j.cell.2020.06.013

15. Li Y, et al. Proteogenomic data and resources for pan-cancer analysis. Cancer Cell. 2023;41:1397–406. doi:10.1016/j.ccell.2023.06.009

16. Karczewski KJ, et al. The mutational constraint spectrum quantified from variation in 141,456 humans. Nature. 2020;581:434–43. doi:10.1038/s41586-020-2308-7

17. Tsherniak A, et al. Defining a cancer dependency map. Cell. 2017;170:564–76. doi:10.1016/j.cell.2017.06.010

18. Giurgiu M, et al. CORUM: the comprehensive resource of mammalian protein complexes— 2019. Nucleic Acids Res. 2019;47:D559–63. doi:10.1093/nar/gky973

19. The UniProt Consortium. UniProt: the Universal Protein Knowledgebase in 2023. Nucleic Acids Res. 2023;51:D523–31. doi:10.1093/nar/gkac1052

20. Bausch-Fluck D, et al. A mass spectrometric-derived cell surface protein atlas. PLoS One. 2015;10:e0121314. doi:10.1371/journal.pone.0121314

21. CZ CELLxGENE Discover: a single-cell data platform for scalable exploration, analysis and modeling of aggregated data. Nucleic Acids Res. 2025;53:D886–900. doi:10.1093/nar/gkae1142

22. Uhlén M, et al. Tissue-based map of the human proteome. Science. 2015;347:1260419. doi:10.1126/science.1260419

23. Karlsson M, et al. A single-cell type transcriptomics map of human tissues. Sci Adv. 2021;7:eabh2169. doi:10.1126/sciadv.abh2169

24. Loyfer N, et al. A DNA methylation atlas of normal human cell types. Nature. 2023;613:355–64. doi:10.1038/s41586-022-05580-6

25. Corces MR, et al. The chromatin accessibility landscape of primary human cancers. Science. 2018;362:eaav1898. doi:10.1126/science.aav1898

26. GTEx Consortium. The GTEx Consortium atlas of genetic regulatory effects across human tissues. Science. 2020;369:1318–30. doi:10.1126/science.aaz1776

27. Chen T, Guestrin C. XGBoost: a scalable tree boosting system. In: Proc. 22nd ACM SIGKDD. 2016:785–94. doi:10.1145/2939672.2939785

28. Efron B, Morris C. Data analysis using Stein’s estimator and its generalizations. J Am Stat Assoc. 1975;70:311–9. doi:10.1080/01621459.1975.10479864

